# Clinician-Informed Feature Engineering Improves Machine Learning Assignment of Molecular Endotypes in the Intensive Care Unit

**DOI:** 10.64898/2026.04.06.26350248

**Authors:** Benjamin J Sines, Robert S Hagan, Xi Jiang, Ella Pavlechko, Scott McClain, Xin Hunt, Julia Florou-Moreno, Jake Acquadro, Gabriel Risa, Varun Valsaraj, Jonathan C Schisler, Matthew C Wolfgang

## Abstract

**Objective:** To develop a workflow that transforms electronic health record data into machine learning-ready features for molecular endotype assignment and to evaluate whether clinician-informed feature engineering improves model performance and interpretability.

**Materials and Methods:** We developed parallel clinician-informed and clinician-agnostic feature engineering pipelines to prepare raw EHR data from mechanically ventilated patients with respiratory failure. Molecular endotype labels derived from paired deep lung and blood profiling of subjects with acute lung injury were used to train candidate machine learning classifiers. Champion models from each pipeline were compared on predefined performance metrics.

**Results:** Bayesian network classifiers were the top-performing models in both pipelines. The clinician-informed pipeline generated fewer features than the clinician-agnostic pipeline (645 vs 1,127) and produced a lower misclassification rate in the final Bayesian network model (0.047 vs 0.14). In an independent cohort of subjects with acute lung injury, the clinician-informed model better distinguished corticosteroid-responsive from non-responsive subgroups.

**Discussion:** Clinical context improved feature engineering efficiency, model interpretability, and classification performance. These findings support the integration of domain expertise into machine learning workflows intended for critical care implementation.

**Conclusions:** Clinician-informed feature engineering can simplify machine learning models while improving performance and preserving clinical relevance. AI tools developed for healthcare should incorporate subject matter expertise early in the feature engineering and analytic workflow.

## BACKGROUND AND SIGNIFICANCE

Advanced machine learning (ML) that leverages data from the Electronic Health Record (EHR) promises enhanced diagnostic, therapeutic, and prognostic decision making in healthcare. This potential is particularly relevant in data-rich, highly monitored clinical environments such as intensive care units. The volume of clinically generated data has grown tremendously since the widespread adoption and integration of electronic health records and is now large enough that applications of machine learning can draw new insights into disease and clinical care.^1^ While emerging algorithmic approaches can identify patterns and draw inferences that elude clinicians, model performance remains highly dependent on feature engineering and on interpretation within the appropriate clinical context.^2–6^ Like other diagnostic tools, machine learning algorithms require careful scrutiny, but they may enable rapid assessments using near real-time clinical data at a scale beyond unaided human interpretation.

Despite rapid advances in ML, clinical adoption in intensive care has been limited by the narrow window for intervention and by insufficient reporting of model performance in clinically actionable terms. Critical care is typified by organ support in which early interventions to bring patient conditions and laboratory parameters to within acceptable norms make a difference in mortality outcomes. Delays in the application of therapies by minutes can translate to increased mortality.^7^ Patient physiological and laboratory values are vastly different in the critically ill as compared to healthy populations and non-critically ill populations.^8–10^ As a result, clinical data used in modeling must be contextualized to the specific pathology and clinical setting of the intended population for model use. For example, timing can have a large impact simply because diagnostics quickly reflect therapeutic intervention. Thus, as the interval between treatment and measurement increases, clinical data may lose predictive or discriminatory value because clinical values progress towards a predefined acceptable range.

The opportunity for improvements in care of the critically ill patient lies in accessing multiple data sources. When made available, clinical data must be prioritized and harmonized for fidelity and completeness for their potential clinical support value. This careful attention to data fidelity is also critical when analytics are used for research purposes and early development of predictive ML models. A key challenge in multi-source clinical data is that duplicate and highly correlated measurements do not add independent information and can instead increase model complexity. These data therefore require dimensionality reduction during feature engineering. This dimensionality reduction enhances model performance and interpretability but requires subject matter expertise and is critical to the overall workflow that can make an analytic approach feasible enough to enter the clinic.^11^

## OBJECTIVES

Incorporating clinical subject matter expertise into analytic approaches is a known value-added function in predictive model development. Feature engineering, for example, can produce models that are easier to interpret without substantially compromising technical performance.^5^ The inclusion of statisticians, clinicians, and diagnostic experts, increases the likelihood that the overall analytic approach will incorporate meaningful domain knowledge. This allows clinically and statistically relevant inputs to be prioritized while less informative variables are removed. This prevents a model from otherwise having important data excluded or dismissed during model development. This is particularly important in approaches that assume more data necessarily yields better results, when in fact excessive feature inclusion may promote overfitting and unreliable predictions.

Using experiential knowledge improves the precision of the mutual information score ranking of features.^12–14^ This is a measure of clinical relevance of features included in models specific to healthcare applications. To our knowledge, the effect of clinician input at the feature engineering stage has not been directly evaluated for ICU machine learning applications. We therefore compared clinician-informed and clinician-agnostic feature engineering pipelines for molecular endotype assignment in mechanically ventilated patients with respiratory failure. We hypothesized that clinician-informed preprocessing would reduce overfitting, improve model interpretability, enhance predictive performance, and offers a path toward the clinical utility of these tools.

## MATERIALS AND METHODS

This study was approved by the institutional review board at the University of North Carolina at Chapel Hill with a waiver of informed consent (IRB 22-3196, January 11, 2023). We followed the Transparent Reporting of a Multivariable Prediction Model for Individual Prognosis or Diagnosis plus Artificial Intelligence (TRIPOD+AI) reporting guidelines for reporting the development and evaluation of the presented models (**Supplement A – Table 1**).^15^

**Table 1:** Baseline Clinical Characteristics of the Development Cohort.

| **Table 1:** Baseline Clinical Characteristics of the Development Cohort |  |
| --- | --- |
| Clinical Characteristics of Development Cohort | N = 43 |
| Age, median (IQR), years | 60.4 [48.5, 67.3] |
| Male Sex, no. (%) | 22 (50.0) |
| Race, no. (%) |  |
| Non-Hispanic white | 24 (54.5) |
| Non-Hispanic black | 17 (38.6) |
| Hispanic | 3 (6.8) |
| BMI, median (IQR) | 31.6 [28.0, 36.1] |
| Hospital Length of Stay, median (IQR), days | 17.0 [12.0, 29.3] |
| ICU Length of Stay, median (IQR), days | 10.0 [8.0, 16.4] |
| Myocardial Infarction, no. (%) | 9 (20.5) |
| Congestive Heart Failure, no. (%) | 8 (18.2) |
| Peripheral Vascular Disease, no. (%) | 5 (11.4) |
| Cerebrovascular Disease, no. (%) | 4 (9.1) |
| Chronic Pulmonary Disease, no. (%) | 10 (22.7) |
| Connective Tissue Disease, no. (%) | 1 (2.3) |
| Diabetes without end-organ complication, no. (%) | 18 (40.9) |
| Diabetes with end-organ complication, no. (%) | 2 (4.5) |
| Chronic Kidney Disease stage 3-5, no. (%) | 4 (9.1) |
| Cirrhosis, no. (%) | 3 (6.8) |
| Charlson Comorbidity Index, median (IQR) | 3 [2, 5] |
| Apache II Score, median (IQR) | 22.0 [18.0, 25.3] |

### Study Population

We utilized clinical and molecular data from patients in the Medical Intensive Care Unit with respiratory failure from COVID-19 and other causes at the University of North Carolina Hospital between April 14, 2020, and February 14, 2021. Subjects were recruited to the study if they required invasive mechanical ventilation for respiratory failure and underwent testing for respiratory viral infections. Deep lung and peripheral blood samples were collected for cytokine profiling and RNA sequencing, which identified two distinct molecular endotypes with differential predicted responses to standard pharmacotherapies.^16^

### Feature Engineering Approach

To evaluate the added value of domain expertise in feature engineering, we developed two parallel feature engineering pipelines (workflows): clinician-informed (CI) and clinician-agnostic (CA). The CA pipeline mirrored the core steps of the CI approach but relied solely on data-driven, objective criteria for feature processing and model development. By conducting a side-by-side comparison of both pipelines, we assessed the contribution of clinical insight to model performance and complexity. Both the CI and CA feature engineering pipelines followed five core steps: data pre-cleaning, data filtering, data preprocessing, feature generation, and postprocessing, as shown in **Figure 1**. The key differences between the two pipelines occur in the preprocessing and feature generation steps, highlighted in green in **Figure 1**. In these steps, the CI pipeline incorporated domain-specific knowledge from clinicians to guide data processing and feature generation based on clinical relevance. Specifically, two clinicians reviewed all available features and prioritized variables based on clinical relevance. Features expected to converge rapidly with treatment, such as oxygen saturation and arterial pressures, were deprioritized relative to measures of organ support, such as fraction of inspired oxygen and vasopressor requirement. When parallel measurements were available, clinicians also imposed a hierarchy of data quality; for example, arterial line blood pressure was prioritized over non-invasive blood pressure, and arterial pH over venous pH. These decisions also informed trend generation, averaging, and binning. The CA pipeline relied solely on statistical methods, treating all features equally without the integration of clinical expertise. Additional details on the pipelines are provided in Error! Reference source not found. and **Figure 2**.

**Figure 1.**
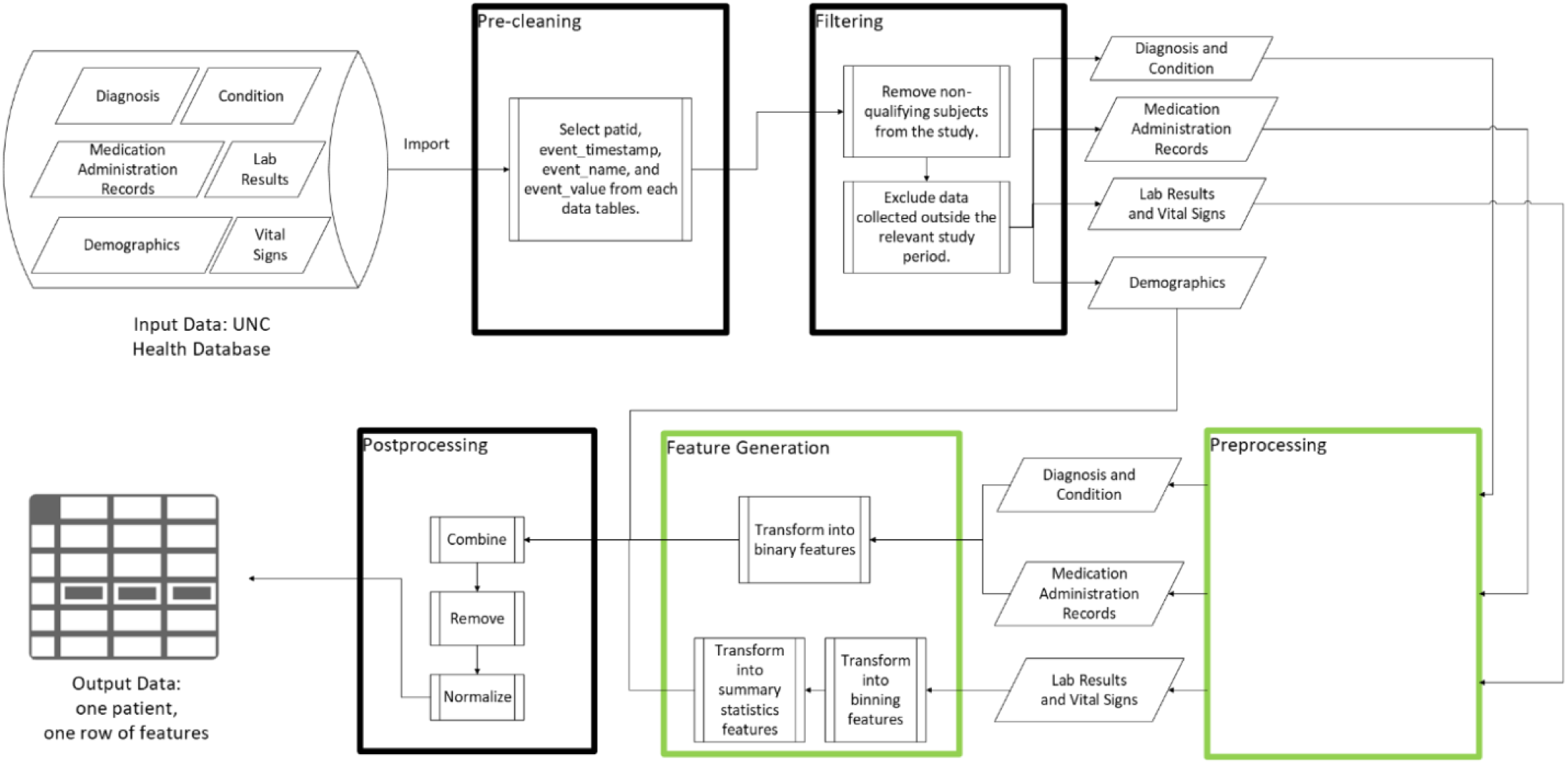
The Overview of Feature Engineering Pipeline(s). Patient-level data was provided in the PCORnet data model. These data flowed through pre-cleaning and filtering steps to identify variables outcomes relevant to the ICU admission of interest. A preprocessing step then condensed redundant diagnoses and medications in preparation for feature generation. Feature generation was subsequently performed via either the clinician-agnostic or clinician-informed pipelines. Finally, data were discretized for machine learning model development.

**Figure 2.**
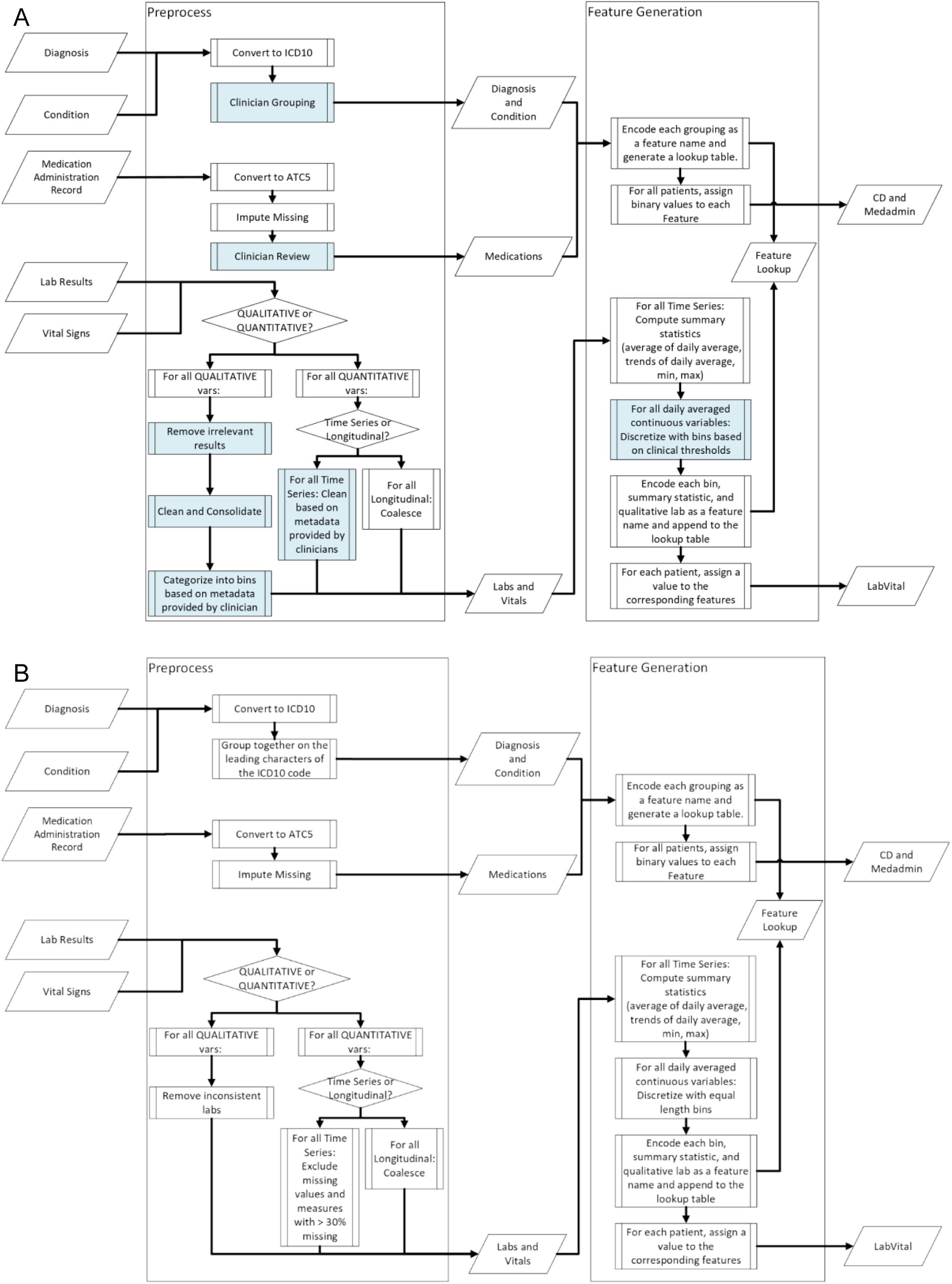
A Detailed View of the Preprocessing and Feature Generation Steps for the **A**) CI and **B**) CA Pipelines. Areas of clinician input is highlighted in blue.

The input to both pipelines consisted of electronic health record data from patients admitted to the medical intensive care unit with respiratory failure. The electronic health record data is archived in the Carolina Data Warehouse for Health at the University of North Carolina. This input data was organized in the PCORnet Common Data Model and included multiple tables capturing diagnoses, clinical conditions, medication administration records, laboratory results, vital signs, and demographics. All tables were linked by a common patient identifier. All data sources underwent the five main steps, except for demographic data (age, sex, and race), which bypassed feature engineering and proceeded directly from filtering to postprocessing.

All data sources were first filtered to retain only measurements from patients who meet the study’s inclusion criteria. Then, the data was further filtered to include only those measurements recorded within the study’s specified time frame. This two-step filtering process ensured the pipeline’s flexibility, allowing it to be tailored to any study of interest. A specific study protocol and further details on possible inclusion criteria and filters are included in the Supplementary Materials.

Standard preliminary data cleaning was performed to assess data completeness and linkages. Data preprocessing varied by tabular data provided in the PCORnet common data model. As part of preprocessing, all condition and diagnosis codes were mapped to the ICD-10 classification system. In the CI pipeline, these codes were further grouped into broader, clinically defined disease categories.

Medication administration records in these data were variably mapped to both NDC codes and RxNorm codes. As a part of the feature engineering pipeline, medication records were initially mapped to RxNorm codes using an API call to the NIH’s RxNav interface (https://rxnav.nlm.nih.gov). In instances where RxNav did not return an RxNorm code, the code was imputed by a text-based comparison of medication names with the DICER algorithm, which is detailed in **Supplement B:** Error! Reference source not found. These codes were then mapped to the hierarchical Anatomical Therapeutic Chemical (ATC) classification for systematized dimensionality reduction.

Laboratory values and vital signs in the intensive care unit frequently have repeated measures with variable intervals and, often, duplicate or highly covariant data streams. For example, prothrombin time and the international normalized ratio (INR) are nearly perfectly collinear, and heart rate is available from both pulse oximeter tracing and continuous telemetry with essentially perfect collinearity. Qualitative laboratory values with multiple labels for dichotomous outcomes were harmonized. Repeated quantitative laboratory values were treated as time series data. Calculated values including body mass index and estimated glomerular filtration rate were imputed using standard formulae when data for inputs were available. Events with high levels of missingness (>0.3) were excluded.

### Derivation of Prediction Models

The predictive modeling process included four main stages: model selection, model training, model evaluation and selection, and model interpretability. All modeling and analysis were performed using SAS Viya 4. Model selection was guided by the ability to perform classification, handle missing data, and provide high interpretability, as well as by the limited dataset of 43 observations. The candidate machine learning models included three tree-based approaches (decision tree, gradient boosting, and random forest), a Bayesian network classifier, and logistic regression.

For model training, hyperparameters were determined through autotuning and evaluated using a macro function that controls randomness and incorporates cross-validation. For each set of hyperparameters, the 43 subjects were divided into 10 groups, so that each subject was used in testing once across the 10 folds of the cross-validation macro. The data for each fold consisted of 90% of patients to train the model then testing with the 10% of remaining patients. The results were consolidated across all folds for evaluation (the full training and evaluation process is summarized in **Figure 3**).

**Figure 3.**
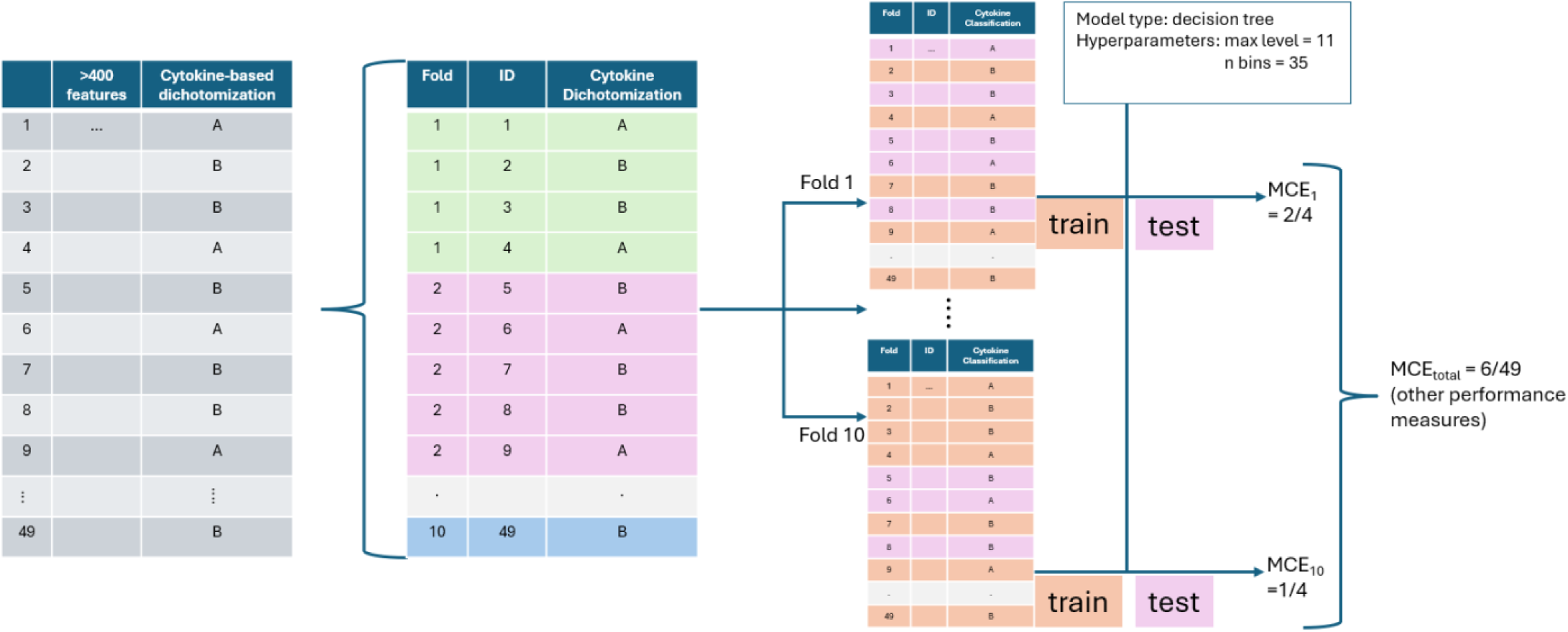
An Overview of the 10-fold Cross-validation Model Training Macro. This training macro was rerun for various hyperparameter choices, over various model architectures, and on CI and CA features.

Model performance was evaluated with classification accuracy and true positive rate across a range of confidence threshold values. Model interpretability was also assessed to understand how each model generates predictions, ensuring not only accuracy but also clinical trust. Finally, we assessed clinician-agnostic vs clinician-informed model capacity to identify corticosteroid responders and non-responders in a large, independent cohort of patients requiring mechanical ventilation for severe COVID-19 pneumonia.

## RESULTS

A total of 88 subjects were recruited to the parent study. Paired cytokine data and RNA sequencing from both deep lung samples and blood were available for 43 subjects, allowing them to be clustered into two distinct molecular endotypes. Clinical variables and demographics for the 43 subjects are displayed in **Table 1**. The data scientists, statisticians, and clinicians were masked to the therapeutic implications of the molecular classes.

### Feature Engineering Output

All available clinical data collected within ±24 hours of enrollment from subjects with an assigned molecular endotype were ingested by the feature engineering pipeline, resulting in 645 clinician-informed features and 1,127 clinician-agnostic features available for modeling.

### Champion Model Selection and Model Performance

The best-performing (champion) models were the CI and CA Bayesian network classifiers. The true positive rate (TPR) and correct classification rate (CCR) from 10-fold cross-validation at a threshold probability of 0.5 are summarized in **Table 2**. The TPR and CCR values above 70% indicated that the decision tree, Bayesian network classifier, and random forest were the top performing models.

**Table 2:** The 10-fold predictive modeling results using the features generated on Day –1 and 0.

|  |  |  |  |  |
| --- | --- | --- | --- | --- |
|  | CI True Positive Rate | CA True Positive Rate | CI Correct Classification Rate (%) | CA Correct Classification Rate (%) |
| Decision Tree | 4/10 (40) | 5/10 (50) | 32/43 (74.4) | 33/43 (76.7) |
| Bayesian network | 8/10 (80) | 7/10 (70) | 35/43 (81.4) | 33/43 (76.7) |
| Random forest | 0/10 (0) | 0/10 (0) | 33/43 (76.7) | 33/43 (76.7) |
| Gradient boosting tree | 0/10 (0) | 1/10 (10) | 32/43 (74.4) | 25/43 (58.1) |
| Logistic Regression | 4/10 (40) | 3/10 (30) | 22/43 (51.2) | 24/43 (55.8) |

The Bayesian network classifiers and decision trees remained the best performing models for class predictor confidence. Varying the predictive confidence minimum threshold (**Figure 4A**) demonstrated that at thresholds above 0.6 the Bayesian network classifiers and random forest models had a >70% CCR. However, at thresholds above 0.75 the random forest models classified fewer than 60% of the population (**Figure 4B)**. As a result, we removed the random forest model from consideration. **Figure 4** also suggests that logistic regression and gradient boosting classified a relatively large portion of the cohort; however, because their accuracy was lower, these models were not considered further.

**Figure 4.**
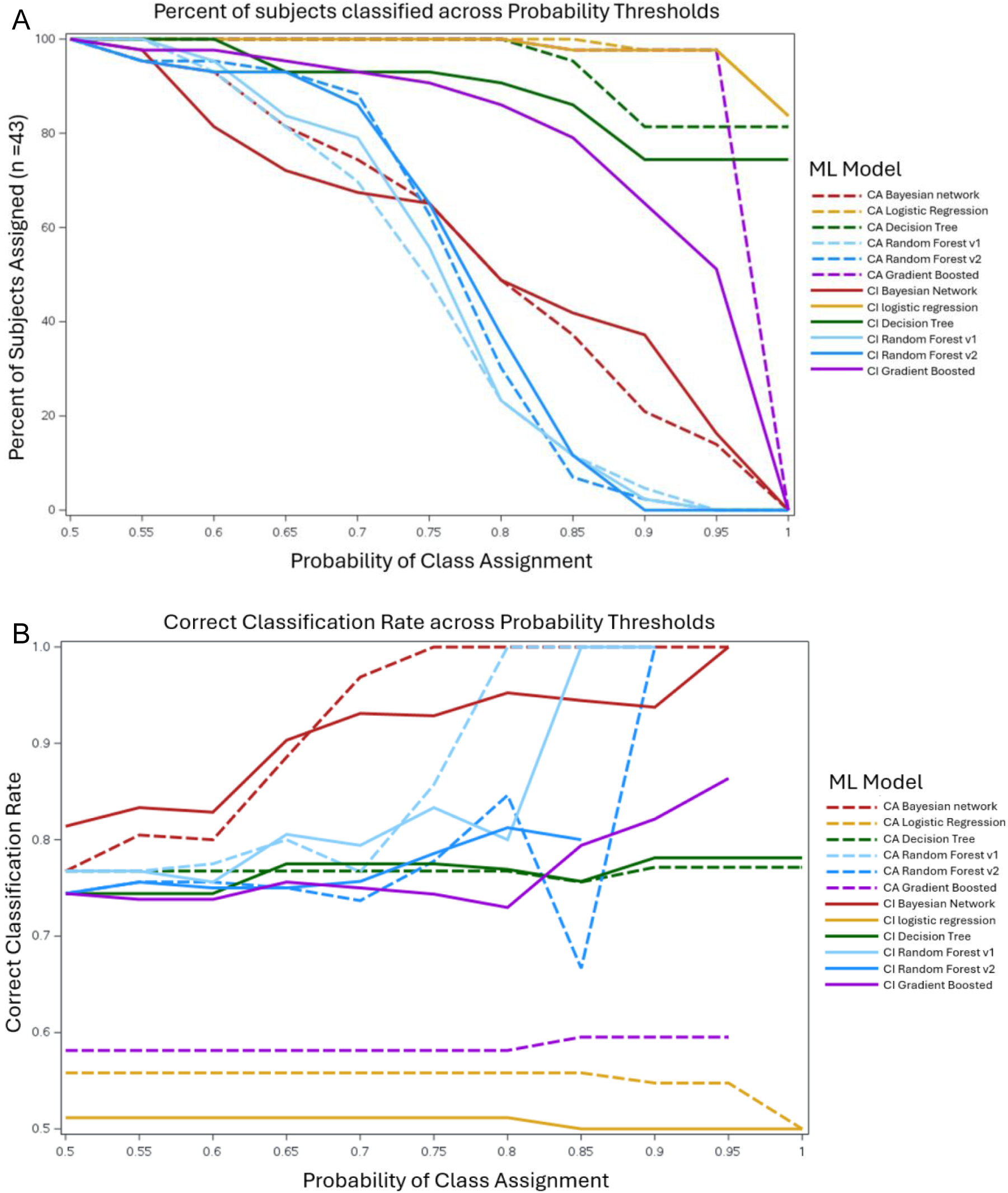
Machine Learning Model Performance Across Probability of Class Assignment. **A**) percent of subjects classifiable at a priori probability thresholds. **B**) Correct classification rate of classified subjects at varying thresholds.

To identify the top performing model among the remaining candidates, we focused on maximizing CCR while also preserving the proportion of the population classified across threshold values (**Table 3**). Because the decision trees maintained a high proportion of classified patients across thresholds, we suspected that the models were overly simple, overfit, or influenced by the small training sample size. To confirm, we analyzed the models in each of the 10-folds. We determined that the decision trees were entirely misclassifying endotype A, the minority class, in 4 out of 10 folds. Therefore, to avoid bias, the Bayesian network classifiers were selected as the champion models.

**Table 3.** Comparison of Correct Classification Rate (CCR) and Percentage Predicted across varying thresholds for top-performing models.

| Threshold<br>(predicted<br>probability) | CA Bayesian Net. |  | CA Decision Tree |  | CI Bayesian Net. |  | CI Decision Tree |  |
| --- | --- | --- | --- | --- | --- | --- | --- | --- |
|  | CCR | Percentage<br>Predicted | CCR | Percentage<br>Predicted | CCR | Percentage<br>Predicted | CCR | Percentage<br>Predicted |
| 0.6 | 0.80 | 93.02 | 0.77 | 100.00 | 0.83 | 81.40 | 0.74 | 100.00 |
| 0.7 | 0.97 | 74.42 | 0.77 | 100.00 | 0.93 | 67.44 | 0.78 | 93.02 |
| 0.8 | 1.00 | 48.84 | 0.77 | 100.00 | 0.95 | 48.84 | 0.77 | 90.70 |
| 0.9 | 1.00 | 20.93 | 0.77 | 81.40 | 0.94 | 37.21 | 0.78 | 74.42 |

The final Bayesian network classifiers were trained on all 43 patient observations. The CI Bayesian network classifier had a misclassification rate of 0.047, while the CA network had a misclassification rate of 0.14. The final trained champion models are displayed in Figure 5. To understand how each feature contributes to the classification, we examined the structure of the top performing model, which had explicit and tractable mathematical formulations, detailed in **Supplement C: Clinician-informed Bayesian Network Model**. Specifically, we found that this model incorporated data elements representing multiple vital organ systems that are routinely tracked and supported in critical care, representing multiple axes of critical illness.

**Figure 5.**
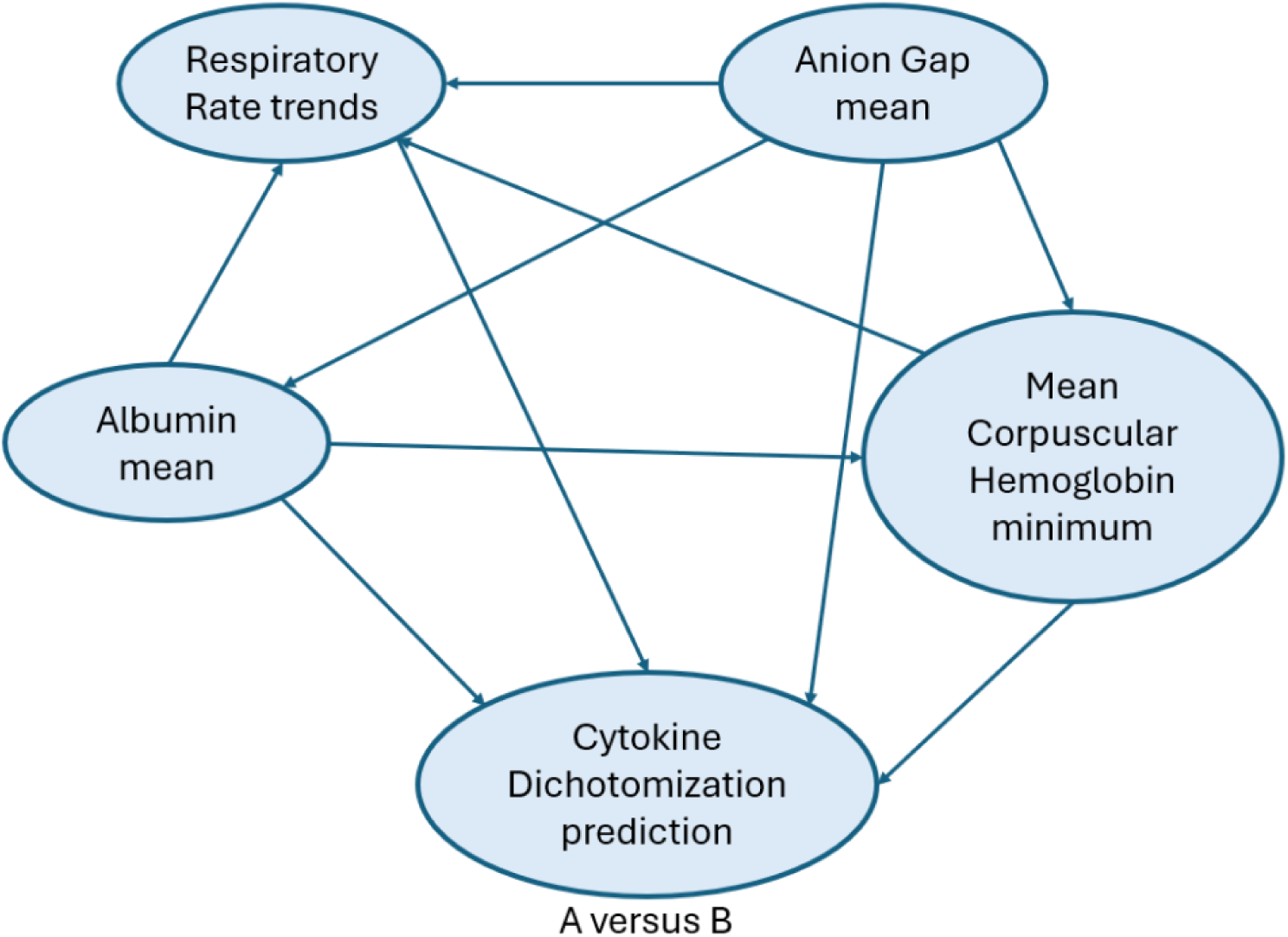
The Champion Model: Bayesian Network Classifier Trained with Clinician-informed Features.

We evaluated the two models in a target trial emulation of corticosteroids for severe COVID-19 pneumonia.^17^ The clinician-informed classifier distinguished subjects with evidence of corticosteroid benefit from those without an apparent clinical response. The odds ratio of 28-day mortality with treatment was 0.62 (95% CI 0.39, 0.99) in clinician-informed Endotype B and 1.15 (95% CI 0.82, 1.61) in clinician-informed Endotype A. On the other hand, the clinician-agnostic model does not separate the emulated target trial population into corticosteroid responders. The odds ratio of 28-day mortality with treatment in clinician-agnostic Endotype B assignment 0.83 (95% CI 0.46, 1.50) versus 1.00 (95% CI 0.73, 1.36) in clinician-agnostic Endotype A assignment **(Table 4)**.

**Table 4:** 28-day Mortality under Treatment with Corticosteroids versus Supportive Care Alone.

|  | Endotype A | Endotype B |
| --- | --- | --- |
| Clinician-Informed Bayesian Network, OR (95% CI) | 1.15<br>(0.82, 1.61) | 0.62<br>(0.39, 0.99) |
| Clinician-Agnostic Bayesian Network, | 1.00<br>(0.73, 1.36) | 0.83<br>(0.46, 1.50) |
| OR (95% CI) |  |  |

## DISCUSSION

In this retrospective cohort, we developed clinician-informed and clinician-agnostic ML models to predict molecular endotype in mechanically ventilated patients with acute lung injury using routinely available clinical features. We integrated tissue-specific, blood-based, and clinical data to derive a feature set suitable for model development. CI feature engineering substantially reduced the input feature space for ML-based diagnostic modeling. Reducing the feature space decreases the risk of overfitting. A reduced feature space also yielded a more interpretable model. This enhanced transparency and trustworthiness are critical for clinician-facing decision support tools.

Machine learning models utilizing the CI feature engineering pipeline performed as well or better than those using the CA pipeline. Although the gain in classification accuracy was modest, the clinician-informed approach prioritized features that were clinically meaningful and mechanistically relevant. For example, trends in respiratory rate reflect illness severity and response to therapy in patients requiring mechanical ventilation for respiratory failure, while the mean anion gap provides another dimension of organ failure and acid-base homeostasis. This yielded predictive models with comparable or improved accuracy despite a smaller feature set. Rather than relying on a brute-force, high-dimensional strategy, CI feature engineering promoted data efficiency. This is especially valuable in real-world clinic settings where data can be sparse, noisy, or expensive to curate, all of which can delay timely care under current approaches.

Moreover, this approach ensured interpretability and clinical relevance. Clinician-informed features were rooted in practice patterns and tacit clinical knowledge, making them inherently more interpretable. This approach helped ensure that the models were not only predictive but also clinically interpretable and actionable. Importantly, by anchoring feature engineering in clinical reasoning, the pipeline avoided the risk of “garbage in, garbage out.” It filtered out irrelevant or misleading signals, allowing the model to focus on predictors aligned with how clinicians assess and manage risk.

### Limitations

This study had some notable limitations. First, real world data generated for the purposes of clinical care has inherent biases and missingness that cannot be readily circumvented with context aware feature engineering. These include provider decision to order laboratory tests, documentation of existing comorbidities, and clinical variability in the timing and choice of medication and procedural interventions in the intensive care unit. Second, the sample size of study subjects used in this analysis was limited to the subjects for whom paired molecular analysis of tracheal aspirate and peripheral blood samples exist. Larger sample sizes and independent prospective validation of these models are critical next step prior to widespread deployment of these clinical models.

## CONCLUSION

This study addressed two related questions: whether clinically relevant patient data can be transformed into a predictive analytic workflow, and whether clinical expertise enhances feature engineering for machine learning models built from real-world ICU data. We implemented and tested a clinician-informed feature engineering research model that transformed clinical data into machine learning–ready features. The key test of the concept was whether the clinician-informed approach could outperform a clinician-agnostic model while using fewer features. Our finding that clinician-informed feature engineering improved model interpretability and accuracy supports the integration of domain experts into this stage of ML development as a practical path toward clinically deployable decision-support tools.

## Supporting information

Supplementary content

## Data Availability

All data produced in the present study are available upon reasonable request to the authors

## Author contributions

Benjamin J Sines (Conceptualization, Data curation, Investigation, Methodology, Project administration, Writing – original draft, Writing-review and editing), Robert S Hagan (Funding, Data curation, Writing-review and editing), Xi Jiang (Data curation, Software, Visualization, Methodology, Writing-review and editing), Ella Pavlechko (Data curation, Software, Visualization, Methodology, Writing-review and editing), Scott McClain (Methodology, Writing-review and editing), Xin Hunt (Data curation, Software, Visualization, Methodology), Julia Florou-Moreno (Data curation, Software), Jake Acquadro (Data curation, Software), Gabriel Risa (Conceptualization, Data curation, Software, Methodology), Varun Valsaraj (Project administration, Software, Methodology, Writing-review and editing), Jonathan C Schisler (Funding, Data curation, Software, Methodology, Writing-review and editing), Matthew C Wolfgang (Funding, Project administration, Conceptualization, Methodology, Writing-review and editing)

