## Supplementary content for "Clinician-Informed Feature Engineering Improves Machine Learning Assignment of Molecular Endotypes in the Intensive Care Unit"

**Conflict of Interest:** The authors have declared that no conflict of interest exists.
Key Words: Machine Learning, Clinical Prediction, ARDS, Critical Care
Word count: 2981

### 1. Supplement B – Feature Engineering

This supplement provides detailed descriptions of the CI and CA Feature Engineering Pipeline. A pipeline overview is provided in **Figure 1**, with a more detailed view of the Preprocessing and Feature Generation steps shown in **Figure 2**.

#### 1.1 Pre-cleaning

Preliminary cleaning was performed on each data source to ensure that every observation included essential elements such as the patient ID, measurement timestamp, measurement name, and measurement value (denoted as either numeric or qualitative).

#### 1.2 Data filtering

All data sources were first filtered to retain only the measurements from patients who meet the study's inclusion criteria. Then, the data was further filtered to include only those measurements recorded within the study's specified time frame. This two-step filtering process ensured the pipeline's flexibility, allowing it to be tailored to any study of interest. In the results section of this paper, we outline a specific study protocol and provided further details on possible inclusion criteria and filters.

#### 1.3 Data preprocessing

Data from various sources were preprocessed using source-specific methods to ensure consistency and prepare the dataset for feature generation, which is described in detail in the subsections that follow.

##### 1.3.1 Diagnosis and Condition

All diagnosis and medical condition data were recorded using ICD-10 codes. ICD-10 codes range in length from 3 to 7 characters and follow a hierarchical structure. The first three characters identify the general category of the diagnosis or condition, providing a broad classification. Characters four through six add greater specificity by detailing the etiology (cause), anatomical location, severity, or other relevant clinical information. When a seventh character is used, it typically serves as an extension to convey additional context, such as the type of encounter (e.g., initial or subsequent) or the healing phase.

In the CA pipeline—developed using statistical and model specific principles with no inclusion of clinically-specific data distributions, hierarchical rankings, or assumptions—we deployed a naive approach to handling ICD-10 codes. This involved preprocessing diagnosis codes by truncating them to the first three characters, discarding any characters beyond that. The assumption behind this simplification was that the general category of the diagnosis, represented by the first three characters, offered a sufficient level of granularity to be relevant to clinical outcomes. For example, diagnosis codes such as *A41.1*, *A41.51*, *A41.89*, and *A41.9* were consolidated to *A41*, which broadly denotes *Other sepsis*. Under this assumption, the specific type of sepsis or causative organism was considered less important—the key consideration was whether the patient was diagnosed with sepsis at all.

For the CI pipeline, in addition to rolling each diagnosis code up to its general category (i.e., the first three characters), we took an additional step of organizing these categories into broader, clinician-defined disease groupings. This added layer of data structure reflected clinical relevance and helped align the data more closely with real-world medical understanding. For example, all categories from *A30–A49* were grouped under *Other bacterial diseases*. This is an important differentiation from how the CA pipeline would group the same diagnosis code, and to see a direct comparison of the two, please refer to Example 1 in **Supplementary Table 2**.

**Supplementary Table 2.** Machine Learning Feature Processing Examples: Clinician-informed (CI) and Clinician-agnostic (CA).

| Example | Step | EHR Data Scenario | Description/Context | CI preprocessing result | CA preprocessing result |
|---|---|---|---|---|---|
| 1 | 1.3.1 Diagnosis and Condition | ICD-10 codes: A41.1, A41.51, A41.89, and A41.9 | Diagnosis codes relating to sepsis | consolidate to the variable A30-A49, which denotes *Other bacterial diseases.* | consolidate to the variable A41, which denotes *Other sepsis* |
| 2 | 1.3.2 Medication Administration Record | ATC codes: H02AB02 and H02AB04 | ATC code referring to dexamethasone and methylprednisolone, two treatments of interest for COVID19 | consolidate to the variable H02A, which denotes *Corticosteroids for systemic use, plain* | consolidate to the variable H02AB, which denotes *Glucocorticoids* |

| | | | | | |
|---|---|---|---|---|---|
| 3 | 1.3.3 Lab Result and Vital Signs | “SARS-CoV-2 PCR” and “SARS-CoV-2 NAA” | Both labs record qualitative results such as "NEGATIVE," "POSITIVE," "DETECTED," and "NOT DETECTED." | 1. consolidate to a 'SARS-CoV-2' variable.<br>2. Mapping "NEGATIVE" and "NOT DETECTED" to a single “negative” category, and "POSITIVE" and "DETECTED" to a “positive” category | 1. consolidate to a 'SARS-CoV-2' variable.<br>2. "NEGATIVE," "POSITIVE," "DETECTED," and "NOT DETECTED," are treated as four separate categories |
| 4 | 1.3.3 Lab Result and Vital Signs | "Bilirubin, UA" | This is an ordinal-type lab with results such as "1+," "2+," "3+," "LARGE," "MODERATE," "SMALL," "NEGATIVE," and "OT", | maps *"NEGATIVE"* and *"OT"* to the “**negative”** group, while all others are categorized as "positive" | |
| 5 | 1.3.3 Lab Result and Vital Signs | supplemental oxygen devices delivering less than 21%, or more than 100% FiO2 | extreme and physiologically implausible values | record excluded due to uncertainty about the intended value | no action |
| 6 | 1.3.3 Lab Result and Vital Signs | heart rate of 9 bpm | extreme and physiologically implausible values | record excluded due to uncertainty about the intended value | no action |
| 7 | 1.3.3 Lab Result and Vital Signs | Creatine Kinase of >250,000 | a laboratory result reported as exceeding the upper limit of detection | correct the record to the upper detection limit | no action |
| 8 | 1.3.3 Lab Result and Vital Signs | Prothrombin time ("PT") and International Normalized Ratio ("INR") | PT is less informative than INR | eliminate the redundant variable | both variables are included |
| 9 | 1.3.3 Lab Result and Vital Signs | "blood pH" and "serum bicarbonate levels" | the same signal under different names | eliminate the redundant variable | both variables are included |
| 10 | 1.3.3 Lab Result and Vital Signs | Sepsis Score or Fall Risk Score | Values are derived and subjective | eliminate the unreliable variable | both variables are included |
| 11 | 1.3.3 Lab Result and Vital Signs | Missing values for total parenteral or enteral nutrition | these values are only recorded when an order is placed. If no such order exists, the data is structurally missing (also referred to as censored), not | record is interpreted as zero | variable is excluded if there is 30% missingness across subjects |

| | | | | | |
|---|---|---|---|---|---|
| | | | simply absent due to error | | |
| 12 | 1.3.3 Lab Result and Vital Signs | Missing values for estimated glomerular filtration rate ("eGFR") | | value is derived from creatinine, age, and gender | variable is excluded if there is 30% missingness across subjects |
| 13 | 1.3.3 Lab Result and Vital Signs | “Glucose”, “Glucose Whole Blood”, or “Glucose, POC” | multiple glucose-related fields | consolidate into a single Glucose variable | no action |
| 14 | 1.3.3 Lab Result and Vital Signs | Missing values for “$pCO_2$, Arterial” | | imputed using “$pCO_2$, Venous” with an appropriate offset (e.g., ±5 mmHg), reflecting physiological differences between sample types | variable is excluded if there is 30% missingness across subjects |
| 15 | 1.3.3 Lab Result and Vital Signs | "Protein, UA" | these laboratory measures are recorded in both numeric and character formats. | character entries (e.g., "*Negative*" or "*Trace*") were mapped to numerical values—such as *0* for "*Negative*" and *5* for "*Trace*"—and used to impute missing numeric data accordingly | |

1.3.2 Medication Administration Records

The medication administration information was recorded using either National Drug Code (NDC) or RxNorm codes, in addition to a column indicating which coding system the data entry uses, and the medication name. The NDC code system is maintained by the FDA and assigns an identifier for each drug product. This means drugs with the same active component, but different brand names may be given different NDC codes. On the other hand, the RxNorm system identifies drugs by the active ingredients, strength, and dose. Mapping NDC codes to RxNorm is thus a many-to-one mapping, so this action was the first step of both pipelines. The mapping was performed using an API call to the NIH’s RxNav interface (https://rxnav.nlm.nih.gov). We additionally captured the active status flag associated with the returned RxNorm concept.

To facilitate grouping of medications by their therapeutic use, RxNorm codes were converted to Anatomical Therapeutic Chemic (ATC) codes. Each ATC code follows the format A##AA## where each letter and number reflects a different level of classification. For example, dexamethasone as a treatment for COVID-19 has an ATC code of H02AB02 which reflects the classifications in **Supplementary Table 3**.

**Supplementary Table 3:** Anatomical Therapeutic Chemical Classification System Hierarchy Example.

| Level | ATC code | ATC Classification |
|---|---|---|
| **1** | **H** | **Systemic Hormonal Preparations (excluding sex hormones and insulins)** |
| **2** | **H02** | **Corticosteroids for systemic use** |
| **3** | **H02A** | **Corticosteroids for systemic use, plain** |
| **4** | **H02AB** | **Glucocorticoids** |
| **5** | **H02AB02** | **dexamethasone** |

In the CA pipeline, we truncated all ATC codes to level 4 (5-digits) for grouping after mapping the RxNorm to ATC. This placed drugs like dexamethasone (ATC = H02AB02) and methylprednisolone (ATC = H02AB04) into the same group, as seen in Example 2 in **Supplementary Table 2**. In several cases the API call to RxNav did not return an ATC code. To overcome this obstacle, we imputed the missing ATC codes by comparing the medication names. If a medication name exactly matched another with a known ATC then the known ATC was utilized. This was often the case when one medication record had an active RxNorm while another medication with the same name had an inactive RxNorm, so the API call returned a value for the former but not the latter.

However, in the case where no medication names were an exact match we performed a two-phase calculation of similarity between the medications with unknown and known ATC codes, which we refered to as the DICER algorithm. For the first phase, we shortened each medication's name to its active component and found the best matches. Let *U* be the dataset of active components with unknown ATC and *K* be the dataset of active components with known ATC. Then we computed

$$DICE(u,k) = \frac{2|u \cap k|}{|u| + |k|}, \qquad u \in U, k \in K.$$

Here $|\cdot|$ refers to the length of the string in characters and $\cap$ is the overlapping words in the strings. For a pair $u_1 \in U, k_1 \in K$ if $DICE(u_1,k_1) \geq 0.9$ and $\mathrm{DICE}(u_1,k_1) = \max_{\mathrm{u \in U, k \in K}}\{\mathrm{DICE(u,k)}\}$ then the ATC code of $u_1$ is imputed from the ATC code of $k_1$. However, most active components matched with several possible $\mathrm{k}_1 \in K$, returning many ATC codes. As an example, ‘SODIUM CHLORIDE 0.9% INTRAVENOUS PIGGYBACK’ had unknown ATC and its active component ‘SODIUM CHLORIDE’ matched with the following:

| Known ATC4 | Matched medication name |
|---|---|
| A12CA | SODIUM CHLORIDE 1 GRAM TABLET |
| B05CB | SODIUM CHLORIDE 0.9 % IRRIGATION SOLUTION |
| B05XA | SODIUM CHLORIDE 0.9 % INTRAVENOUS SOLUTION |
| R01AX | SODIUM CHLORIDE 0.9 % FOR NEBULIZATION |
| S01XA | SODIUM CHLORIDE 5 % EYE DROPS |

For those active components with multiple possible ATC codes, we completed the second phase of matching using the entire medication name. That is, for the datasets $\widetilde{U}, \widetilde{K}$ containing the full names of the medications with unknown ATC and known ATC from phase 1 respectively, then a pair $u_2 \in \widetilde{U}, k_2 \in \widetilde{K}$ is matched when $DICE(u_2, k_2) \geq 0.5$ and $\mathrm{DICE}(u_2, k_2) = \max_{u \in \widetilde{U}, k \in \widetilde{K}}\{\mathrm{DICE}(u, k)\}$. Thus, in the second phase the medication ‘SODIUM CHLORIDE 0.9% INTRAVENOUS PIGGYBACK’ was matched with ‘SODIUM CHLORIDE 0.9% INTRAVENOUS SOLUTION’ and given the ATC code B05XA. Using this method means not all medications were able to be mapped to an ATC code, and in the CA pipeline the records that remained unmatched were discarded.

The CI pipeline largely adhered to the same methodology, with two notable deviations. The first is that we truncated the ATC code to level 3 (4 digits). This lower level of grouping maintained the pharmacological purpose sought out by clinicians while removing the superfluous chemical subgroup. The second difference is that a clinician reviewed any conflicting ATC codes that arose at the end of imputation, selected the preferred codes, and assigned an ATC code for the records that remained unmatched using a lookup table (following step 1b, 8d). This ensured all drugs of potential importance for the study were utilized in the model.

1.3.3 Lab Result and Vital Signs

All vital sign measurements were numerical, whereas lab result values can be either numeric or qualitative. In the PCORnet CDM, lab results were stored using two separate fields: RESULT_NUM for numeric values and RESULT_QUAL for character-based (qualitative) values. While most labs were recorded in one format depending on the nature of the test, some labs—such as "Protein, UA"—could appear in both formats (e.g., numeric values like *30*, *100*, *300*, or *500*, in mg/dL, as well as qualitative values like *"NEGATIVE"* or *"TRACE"*).

To handle this variability, both pipelines began with the same processing step: classifying each lab test as either quantitative or qualitative. This classification was based on whether any numeric values were recorded for the lab—if none are found, the test was considered qualitative. As a final clean-up step for the CA pipeline, qualitative lab entries with multiple conflicting results at the same timestamp were treated as erroneous and removed.

For qualitative labs, the CI pipeline included an additional processing step that standardized results by collapsing them into clinically meaningful binary categories—**"**positive**"** and **"**negative**."** It also consolidated related lab tests into unified groups, based on domain-expert knowledge outlined in a reference metadata sheet. Two sample scenarios are laid out in Examples 3 and 4 in **Supplementary Table 2**.

Repeated quantitative labs were technically treated as time series data. However, certain measures (e.g., weight, height and BMI) required additional processing for study-specific times spans less than a week. To ensure accuracy, the CA and CI pipelines both categorized these data as either time series or longitudinal. For longitudinal variables like height, weight, and BMI, only the earliest available records within the specified study window were retained. Missing BMI values were calculated from recorded height and weight using the standard deterministic BMI formula, requiring no clinical inference.

Lastly, data cleaning focused on time series data, such as lab results and vital signs, to resolve issues involving duplicates, missing values, inconsistencies, and errors. In the CA pipeline, any event with more than 30% missingness across subjects was excluded to maintain data integrity. In contrast, the CI pipeline followed domain-specific rules defined in a metadata sheet to guide the handling of data quality issues more precisely. The specific columns utilized and their roles in the cleaning process are detailed in the following paragraphs.

The physiologically possible minimum and maximum values represented the lowest and highest values that a human body is known to present for vital signs and common laboratory results. Occasionally, erroneous data might be entered at the bedside—such as a dropped or extra digit in a heart rate measure or entering a valid measurement into the wrong column; what was otherwise considered an outlier. This sometimes resulted in implausible global minima or maxima. It is essential to handle these inaccuracies carefully through exclusion or correction using realistic local ranges. Some values were adjusted to physiologically plausible local extrema (Example 7 in **Supplementary Table 2**), while others were excluded if the true value was deemed indeterminable (Examples 5 and 6 in **Supplementary Table 2**).

Additionally, for erroneous data management, there were four clinician-evaluated metadata columns that act as a filtering mechanism to eliminate unreliable or redundant entries. Several EHR scenarios with erroneous data are shown in Examples 8-10 in **Supplementary Table 2**. Entries lacking values in all four metadata values were automatically removed from the dataset.

A separate metadata column governed the handling of missing data, guiding manual processing steps through workflows tailored to each measurement. The strategy for addressing missingness was contingent upon the clinical context and the nature of the data element. For example, missing values were either interpreted as zero or imputed using statistical methods, such as mean, median, or multiple imputation by chained equations (MICE), depending on clinical context. Examples 11-15 in **Supplementary Table 2** lay out various scenarios for handling missingness in lab results and vital sign data.

#### 1.4 Feature generation

In the feature generation step of the pipeline, we transformed clean clinical data into structured features tailored to the clinical context of each variable. This included the techniques binary encoding, binning, and summary statistics, all designed to align the data into a format suitable for machine learning algorithms. Binary encoding reduced the number of new columns, helping avoid curse of dimensionality which can lead to overfitting. In addition, tree-based models like decision trees can handle binary-encoded variables more efficiently than columns with sparse values. Binning transformed continuous numerical variables into discrete intervals or "bins". This transformation improved the interpretability of the models as clinicians often think in ranges than raw numbers. For instance, it is easier to interpret a model that flags risk for patients based on their clinical value is above a threshold. Non-linear relationships are often more effectively captured with binning, which can reveal threshold effects— for example, when patient mortality risk jumps sharply once a variable exceeds a specific value.

1.4.1 Diagnosis, Condition, and Medication Administration Records

Diagnosis, condition, and medication administration records are all categorical or event-based data, which were transformed into binary features (e.g., "has diabetes" → 1, "no diabetes" → 0). This binary encoding process was consistent across both pipelines. However, the categorization of these variables differed as a result of the earlier preprocessing step. In the CA pipeline, categories were based on industry-standard groupings such as ICD-10 general categories and ATC4 classifications. In contrast, the CI pipeline applied broader, clinician-defined groupings that reflect higher-level clinical concepts to clean and curate the variables.

1.4.2 Lab Result and Vital Signs

In the CA pipeline, features based on summary statistics of the data were first generated, including the minimum, maximum, and average values. To capture temporal dynamics, trends were estimated by regressing the daily average of each measurement on the corresponding day. This approach accounted for the fact that lab results and vital signs were recorded at varying time intervals and considered only changes across 24-hour periods as a standardized method for aligning temporal resolution. Additionally, to more comprehensively represent the distribution of measurements, binning was applied. Specifically, the probability that the daily average falls within each of five equally spaced bins—determined by the observed range of each variable—was calculated.

On the other hand, the CI pipeline was guided by clinical expertise, as captured in a metadata file. This metadata provided detailed instructions for each lab test or vital sign (identified by EVENT_NAME), specifying how to generate summary statistics (FE_NOTE) and how to apply binning (e.g., N_EDGES, EDGE_0, …, EDGE_7, MISSING_IS_SPEICAL_BIN).

Rather than applying a uniform set of summary statistics to all variables, the FE_NOTE column assigned each variable into one of three processing types: “*average*”, “*min, max, average, trends*”, and “*daily aggregate*”. For the “*average*” type, only the overall average of all measurements across the entire study period was computed. The “*min, max, average, trends*” type included the absolute minimum and maximum values of all measurements, as well as the average and trend computed from daily averages. For the “*daily aggregate*” type, measurements were first summed within each 24-hour period, and then the minimum, maximum, average, and trend were calculated from these daily aggregates. These categories were designed to align with the way each variable was recorded and to preserve relevant physiological meaning. Specifically, some measurements represented discrete events that can be summed over a 24-hour period—for example, total urine output, then “*daily aggregate*” was assigned. Others involved repeated measurements throughout the day and required averaging, trend analysis, or variability assessment. This included most lab and vital sign measurements, such as systolic and diastolic blood pressure, and hematocrit (HCT). For these variables, then “*min, max, average, trends*” was assigned. Additionally, several electrolyte levels—like sodium, potassium, and bicarbonate—were often influenced by pharmacologic interventions. These labs were frequently remeasured after treatment, and their daily averages tended to regress toward clinical targets. Special handling was required for these measures, such as the first recorded value in the calendar day, to capture physiologic deviation from the norm. Pre-defined time interval lengths were determined based on clinical experience as well the practical limitations in capturing real-time observations of changes over time. For example, in patients who are critically ill with respiratory failure, clinicians expect rapid changes in oxygen saturation and supplemental oxygen requirement over the course of hours. Many laboratory measures, such as white blood cell count, would be expected to significantly change only over the course of 24-hour periods. However, the model did not use time units more granular than 24 hours due to variability in data availability and the need for a standardized alignment across all variables. Many lab and vital sign measurements were not recorded frequently or consistently enough to support finer time resolution, making the 24-hour window a practical and clinically meaningful compromise.

The next step in the CI pipeline was the binning process, which handled extreme values through appropriate discretization techniques to capture the underlying distribution of each variable. This process was guided by a metadata sheet that included the columns N_EDGES, EDGE_0 through EDGE_7, and MISSING_IS_SPECIAL_BIN. These fields supported automated, yet clinician-informed binning tailored to both the data distribution and the physiological significance of each measurement. N_EDGES defined the number of bin edges, effectively specifying how many bins span the physiologically plausible range for a given variable. The columns EDGE_0 to EDGE_7 listed the specific bin boundaries, allowing for up to seven edges and six bins—sufficient to accommodate variables with complex distributions. The MISSING_IS_SPECIAL_BIN flag indicated whether missing values should be handled as a separate, dedicated bin.

Bin definitions were shaped by a combination of empirical data distribution, clinical knowledge of physiology, and expectations based on illness severity in the patient population. Different strategies were applied to address extreme values at both ends of the measurement range. For instance, some lab values might appear at magnitudes 10 to 100 times higher than adjacent values—yet still represent true pathological states. Rather than transforming or excluding these outliers, they were included in the highest bin (e.g., creatinine kinase level ranging from 25,000 to >250,000). In cases where measurements spanned orders of magnitude, logarithmic binning was used to preserve clinical relevance across the range. For example, aspartate aminotransferase (AST) and alkaline phosphatase used bin boundaries such as 0, 10, 100, 1,000, and 10,000 to reflect their skewed distribution and meaningful clinical thresholds. In addition, bin boundaries reflected treatment thresholds rather than population-wide norms. For example, instead of using general reference ranges for hemoglobin, a bin threshold was placed at 7 g/dL to align with the standard transfusion trigger in intensive care settings.

1.5 Data postprocessing:

Once all features were generated, postprocessing combined them into a single table, where each patient was represented as one row. Features included diagnosis and conditions (labeled as c_xxx), medication administration records (labeled as m_xxx), qualitative lab results (labeled as l_qual_xx), and quantitative lab tests and vital signs (labeled as lv_num_xxx). Features were then filtered based on their level of missingness, and all remaining features were normalized to a range of -1 to 1. This normalization ensured that each feature contributes equally to the model's learning process, improving model performance and accuracy.

Dimensionality reduction using Principal Component Analysis (PCA) was bypassed to preserve the clinical interpretability of model. Unlike PCA, which transforms variables into abstract components, retaining original features ensures that model output remain mapped to recognizable physiological markers. Additionally, correlation-based feature selection was not utilized; instead, we replied on inherent feature selection mechanisms of the chosen machine learning algorithms to explore the high-dimensional feature space. This approach allowed these models to account for complex feature interactions that might be obscured by pre-filtering. The normalization and preprocessing procedures detailed previously ensured an unbiased performance comparison between the CI and the CA models. In clinical analytics, the ability to trace predictions back to specific variables is essential for transparency and clinician trust, a level of explainability that dimensionality reduction techniques like PCA would fundamentally diminish.

**2. Supplement C:** Clinician-informed Bayesian Network Model Providing Predictive Probability for Endotype A and B.

$$P(A) = \frac{\exp(C_A)}{\exp(C_A) + \exp(C_B)}$$

$$P(B) = \frac{\exp(C_B)}{\exp(C_A) + \exp\,(C_B)}$$

$$where$$

$$C_A = C(A|Respiratory\ Rate\ Trends) + C(A|Albumin\ mean) + C(A|MCH\ minimum) + C(A|Anion\ Gap\ mean),$$

$$C_B = C(B|Respiratory\ Rate\ Trends) + C(B|Albumin\ mean) + C(B|MCH\ minimum) + C(B|Anion\ Gap\ mean)$$

$$C(A|Respiratory\ Rate\ trends) = \begin{Bmatrix} -2.7081\ if -1 \leq Respiratory\ Rate\ trends\ < -0.0354 \\ -1.0986\ if -0.0354 \leq Respiratory\ Rate\ trends\ < 0.1646 \\ -0.9163\ if\ 0.1646 \leq Respiratory\ Rate\ trends\ < 0.3646 \\ -2.0149\ if\ 0.3646 \leq Respiratory\ Rate\ trends\ \leq 1, \end{Bmatrix}$$

$$C(B|Respiratory\ Rate\ trends) = \begin{Bmatrix} -1.6917\ if -1 \leq Respiratory\ Rate\ trends < -0.2354 \\ -0.9985\ if -0.2354 \leq Respiratory\ Rate\ trends < -0.0354 \\ -1.1527\ if -0.0354 \leq Respiratory\ Rate\ trends < 0.1656 \\ -2.9444\ if\ 0.1656 \leq Respiratory\ Rate\ trends < 0.3646 \\ -2.5390\ if\ 0.3646 \leq Respiratory\ Rate\ trends \leq 1, \end{Bmatrix}$$

$$C(A|Albumin\ mean) = \begin{Bmatrix} -2.0149\ if -1 \leq Albumin\ mean\ < -0.3474 \\ -2.7081\ if -0.3474 \leq Albumin\ mean\ < -0.1474 \\ -0.5108\ if -0.1474 \leq Albumin\ mean\ < 0.0526 \\ -2.0149\ if\ 0.0526 \leq Albumin\ mean\ \leq 0.2526 \\ -2.7081\ if\ 0.2526 \leq Albumin\ mean \leq 1, \end{Bmatrix}$$

$$C(B|Albumin\ mean) = \begin{cases} -2.9444\ if -1 \leq Albumin\ mean\ < -0.3474 \\ -1.3350\ if -0.3474 \leq Albumin\ mean\ < -0.1474 \\ -1.0726\ if -0.1474 \leq Albumin\ mean\ < 0.0526 \\ -2.0281\ if\ 0.0526 \leq Albumin\ mean\ \leq 0.2526 \\ -1.5581\ if\ 0.2526 \leq Albumin\ mean \leq 1, \end{cases}$$

$$C(A|MCH\ minimum) = \begin{cases} -0.9163\ if -1 \leq MCH\ minimum < -0.1684 \\ -2.7081\ if -0.1684 \leq MCH\ minimum < 0.0316 \\ -2.0149\ if\ 0.0316 \leq MCH\ minimum < 0.4316 \\ -1.3218\ if\ 0.4316 \leq MCH\ minimum \leq 1, \end{cases}$$

$$C(B|MCH\ minimum) = \begin{cases} -1.4404\ if -1 \leq MCH\ minimum < -0.1684 \\ -0.9985\ if -0.1684 \leq MCH\ minimum < 0.0316 \\ -1.4404\ if\ 0.0316 \leq MCH\ minimum < 0.2316 \\ -2.0281\ if\ 0.2316 \leq MCH\ minimum < 0.4316 \\ -3.6376\ if\ 0.4316 \leq MCH\ minimum\ \leq 1, \end{cases}$$

$$C(A|Anion\ Gap\ mean) = \begin{cases} -2.7081\ if -1 \leq Anion\ Gap\ mean < -0.3093 \\ -0.7621\ if -0.3093 \leq Anion\ Gap\ mean < -0.1093 \\ -1.3218\ if -0.1093 \leq Anion\ Gap\ mean < 0.0907 \\ -2.7081\ if\ 0.0907 \leq Anion\ Gap\ mean < 0.2907 \\ -2.0149\ if\ 0.2907 \leq Anion\ Gap\ mean \leq 1, and \end{cases}$$

$$C(B|Anion\ Gap\ mean) = \begin{cases} -1.6917\ if -1 \leq Anion\ Gap\ mean < -0.3093 \\ -2.0281\ if -0.3093 \leq Anion\ Gap\ mean < -0.1093 \\ -1.4404\ if -0.1093 \leq Anion\ Gap\ mean < 0.0907 \\ -1.3350\ if\ 0.0907 \leq Anion\ Gap\ mean < 0.2907 \\ -1.6917\ if\ 0.2907 \leq Anion\ Gap\ mean \leq 1, and \end{cases}$$
